# Impact of the COVID-19 pandemic on routine immunization coverage in children under 2 years old in Ontario, Canada: A retrospective cohort study

**DOI:** 10.1101/2021.10.28.21265578

**Authors:** Catherine Ji, Pierre-Philippe Piché-Renaud, Jemisha Apajee, Ellen Stephenson, Milena Forte, Jeremy N. Friedman, Michelle Science, Stanley Zlotkin, Shaun K. Morris, Karen Tu

## Abstract

**Background:** The COVID-19 pandemic has caused a disruption in childhood immunization coverage around the world. This study aimed to determine the change in immunization coverage for children under 2 years old in Ontario, Canada, comparing time periods pre-pandemic to during the pandemic.

**Methods:** We conducted an observational retrospective open cohort study, using primary care electronic medical record data from the University of Toronto Practice-Based Research Network (UTOPIAN) database, from January 2019 to December 2020. Children under 2 years old who had at least 2 visits recorded in UTOPIAN were included. We measured up-to-date (UTD) immunization coverage rates, overall and by type of vaccine (DTaP-IPV-Hib, Pneu-C-13, Rot, Men-C-C, MMR, Var), and on-time immunization coverage rates by age milestone (2, 4, 6, 12, 15 and 18 months). We compared average coverage rates over 3 periods of time: January 2019-March 2020 (T1); March-July 2020 (T2); and August-December 2020 (T3).

**Results:** 12,313 children were included. Overall UTD coverage for all children was 71.0% in T1, dropped by 5.7% (95% CI: -6.2, -5.1) in T2, slightly increased in T3 but remained lower than in T1. MMR vaccine UTD coverage slightly decreased in T2 and T3 by approximately 2%. The largest decreases were seen at ages 15-month and 18-month old, with drops in on-time coverage of 14.7% (95% CI: -18.7, -10.6) and 16.4% (95% CI: -20.0, -12.8) respectively during T2. When stratified by sociodemographic characteristics, no specific subgroup of children was found to have been differentially impacted by the pandemic.

**Conclusion:** Childhood immunization coverage rates for children under 2 years in Ontario decreased significantly during the early period of the COVID-19 pandemic and only partially recovered during the rest of 2020. Public health and educational interventions for providers and parents are needed to ensure adequate catch-up of delayed/missed immunizations to prevent potential outbreaks of vaccine-preventable diseases.

## Introduction

The coronavirus disease 2019 (COVID-19) pandemic and associated public health measures aiming to limit its transmission have had an unprecedented impact on healthcare services at all levels, including on routine childhood immunizations [1-3]. Multiple jurisdictions worldwide have reported significant declines in childhood vaccines administered and immunization coverage in the months following the pandemic declaration [4-12]. Disruptions to routine childhood immunization services leading to decreased coverage are concerning for possible resurgence of vaccine-preventable diseases (VPDs), including measles [13-15].

In response to these concerns, international and Canadian public health and governmental institutions have recommended that routine immunizations are essential health services that should not be deferred, especially the primary series and booster doses for children aged less than two years [16-18]. Some studies have nonetheless reported on parents avoiding medical care facilities, clinics not providing in-person services due to the perceived risk of COVID-19 transmission, and school immunization programs being suspended [19, 20]. There is currently limited data to quantify the impact of the pandemic on pediatric immunization coverage in Canada, and whether the pandemic has impacted access to vaccination differentially for specific groups of children. The objective of this study was to compare the immunization coverage for children under two years old before and during the first waves of the COVID-19 pandemic by using primary care electronic medical records (EMR) data in the province of Ontario.

## Methods

### Study design

We conducted an observational retrospective open cohort study using EMR data from family medicine practices included in the University of Toronto Practice-Based Research Network (UTOPIAN) to assess immunization coverage from January 2019 to December 2020.

### Data source

We used data from the UTOPIAN Data Safe Haven, which is a secure research database comprised of de-identified patient records extracted from EMRs of contributing family medicine practices associated with the University of Toronto, with more than 70% of these family physicians practicing in the Greater Toronto Area (GTA) in Ontario [21]. From the UTOPIAN database, we extracted the sociodemographic characteristics (date of birth, sex and postal code) and immunization records (name of vaccine and date given) of the children included in the study population. Measures of socioeconomic status (SES) and marginalization based on neighborhood income, material deprivation and ethnic concentration quintiles were derived based on the children’s postal code using the Statistics Canada Postal Code Conversion File [22-24].

### Study population

We created an open cohort of children using the following eligibility criteria: 1) were aged 0-2 years old during the study period, 2) had a valid sex and date of birth recorded in their EMR, 3) had at least two visits recorded in their EMR during the study period, with at least one visit after the second week of life to ensure continuity of care.

### Setting

Ontario is Canada’s most populous province, with nearly 40% of the country’s population [25]. Early in the COVID-19 pandemic, Ontario declared a state of emergency on March 17, 2020 and entered its first “lockdown” [26]. As the first wave of the pandemic improved, restrictions were progressively lifted throughout the province with Toronto being one of the last regions to enter the final stage of reopening on July 31, 2020 [27]. Ontario experienced a second wave of COVID-19 cases and deaths from September 2020 to February 2021 [28]. At the end of the study period, Ontario had experienced the second highest number of COVID-19 cases and number of deaths in Canada [29]. Within the province, 30% of all cases have been detected in Toronto and the GTA [28].

In Ontario, immunizations for children under two years old are usually administered by primary care providers, including family physicians, during regular well-baby visits [20, 30] and documented in the children’s medical record. EMR data from family physicians’ clinics has been previously used and validated to study childhood immunization coverage in Ontario [30-32]. Ontario’s Publicly Funded Immunization Schedule [33] recommends six different vaccines in the first 24 months of life (Table 1), including diphtheria, tetanus, pertussis, polio, haemophilus influenzae type b (DTaP-IPV-Hib) vaccine, pneumococcal conjugate (Pneu-C-13) vaccine, rotavirus (Rot) vaccine, meningococcal conjugate (Men-C-C) vaccine, measles, mumps, and rubella (MMR) vaccine, and varicella (Var) vaccine.

**Table 1.** Ontario’s Publicly Funded Immunization Schedule for children under 2 years old.

|  | **Age milestones** |  |  |  |  |  |
| --- | --- | --- | --- | --- | --- | --- |
| **Vaccine** | 2 months | 4 months | 6 months | 12 months | 15 months | 18 months |
| DTaP-IPV-Hib | X | X | X |  |  | X |
| Pneu-C-13 | X | X |  | X |  |  |
| Rot | X | X | X* |  |  |  |
| Men-C-C |  |  |  | X |  |  |
| MMR |  |  |  | X |  |  |
| Var |  |  |  |  | X |  |

*\*The Ontario publicly funded immunization program transitioned from the monovalent Rot-1 vaccine (Rotarix) that required two doses at 2 and 4 months to the pentavalent Rot-5 vaccine (RotaTeq) that requires three doses at 2, 4 and 6 months of age in the summer of 2018 [34].*

### Outcomes

Our study had two primary outcome measures: up-to-date (UTD) immunization coverage and on-time immunization coverage. UTD immunization refers to receiving all recommended vaccines by a certain age or by a specific point in time [35, 36]. On-time immunization refers to receiving the recommended vaccines for a specific age milestone within a 30-day leeway period after the due date [35, 36]. For example, a 12-month-old child should receive Pneu-C-13, Men-C-C and MMR vaccines within 30 days after the recommended age to be considered on-time and should have received all doses of recommended vaccines since birth to be considered UTD (Table 1).

### Statistical analysis

The study population’s sociodemographic characteristics were summarized using frequencies and percentages. **Overall UTD coverage rate** for all children under 2 years old for all vaccines was calculated as the number of children who have received all the recommended vaccines for their age milestone divided by the number of children aged 2 to 24 months old. **UTD coverage rate for each vaccine** (DTaP-IPV-Hib, Pneu-C-13, Rot, Men-C-C, MMR, Var) was calculated as the number of children who have received the recommended number of doses of the specific vaccine for their age milestone divided by the number of children who are eligible to receive the vaccine up to 24 months old. **On-time coverage rate for each age milestone** (2, 4, 6, 12, 15 and 18 months) was calculated as the number of children who have received all recommended vaccines at the target age plus a 30-day leeway period divided by the number of children at the target age.

We calculated weekly (Sunday to Saturday) coverage rates from January 6, 2019 to December 20, 2020, which were the start of the first and last full weeks in 2019 and 2020 respectively. We used linear regression models to compare the average immunization coverage rates over three periods of time: baseline/pre-pandemic period, from January 6, 2019 to March 14, 2020 (T1); early pandemic period during the first wave/lockdown in Ontario, from March 15 to August 1, 2020 (T2); and later pandemic period during the reopening phase after first wave and beginning of second wave, from August 2 to December 26, 2020 (T3). We also examined if the differences in on-time immunization coverage rates over the three time periods were associated with any sociodemographic characteristics by fitting adjusted regression models. Each of these models included time period (to calculate the adjusted difference in immunization rates between time periods), one sociodemographic characteristic (to calculate the baseline difference in immunization rates for each level of the characteristic) and the interaction between time period and the sociodemographic characteristic (to calculate difference in differences). We also computed likelihood ratio statistics to assess the contribution of the interaction term to the regression model. Analyses were performed in R version 3.6.1 and SAS 9.4.

### Ethics approval

The study was approved by the University Health Network (#20-6177) and the University of Toronto Health Sciences Research Ethics Boards (#40300).

## Results

A total of 12,313 children aged 0 to 2 years old were included in our study population. The majority of children lived in an urban region (87.2%) and in a neighborhood of the most ethnically diverse quintile (33.4%). They were otherwise almost equally distributed across birth year, sex, neighborhood income and material deprivation quintiles (Table 2). They had a median of 9 (interquartile range: 6-13) visits with their family physician during the study period, which corresponds to the number of recommended routine care visits for young children [37].

**Table 2.**
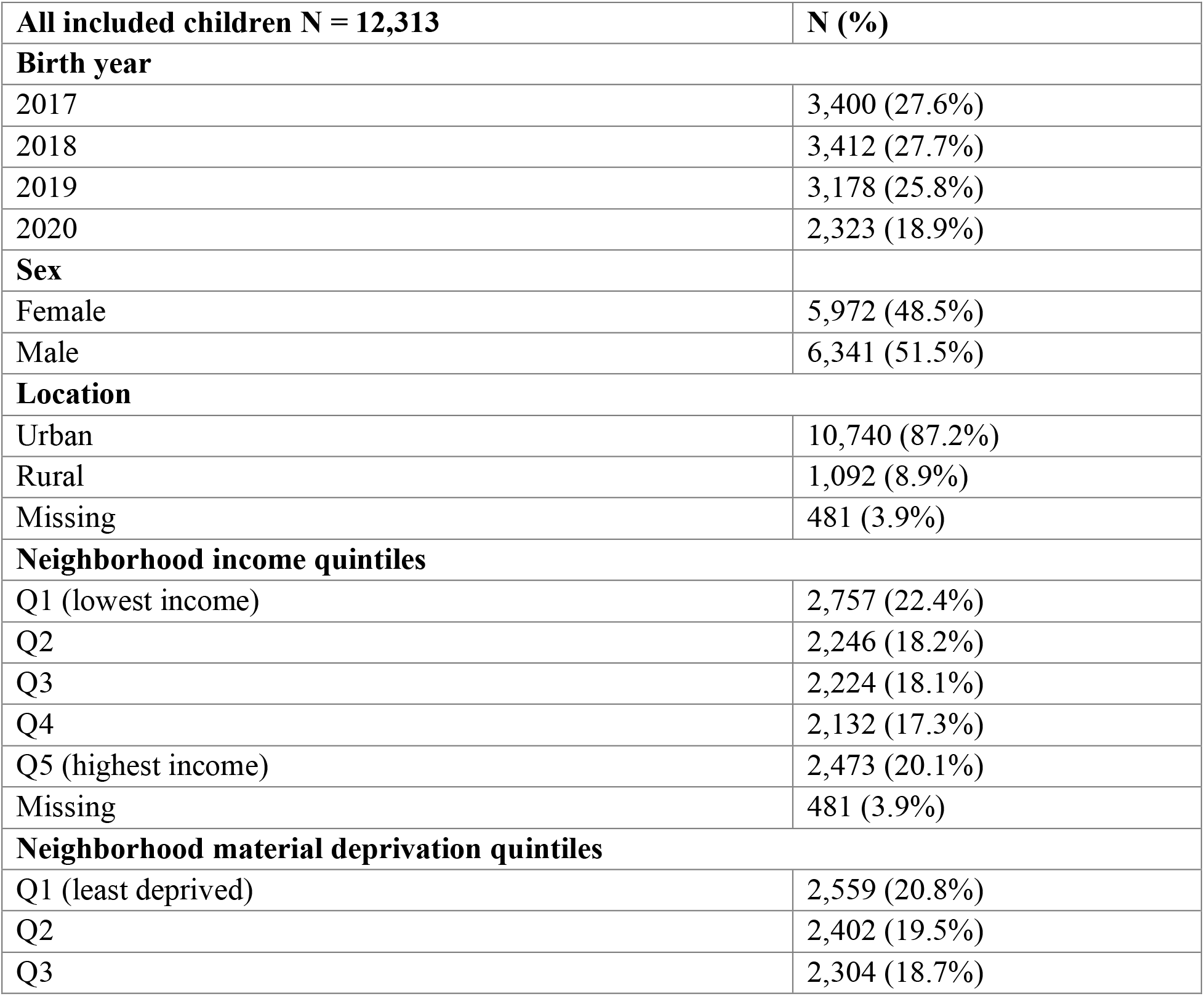
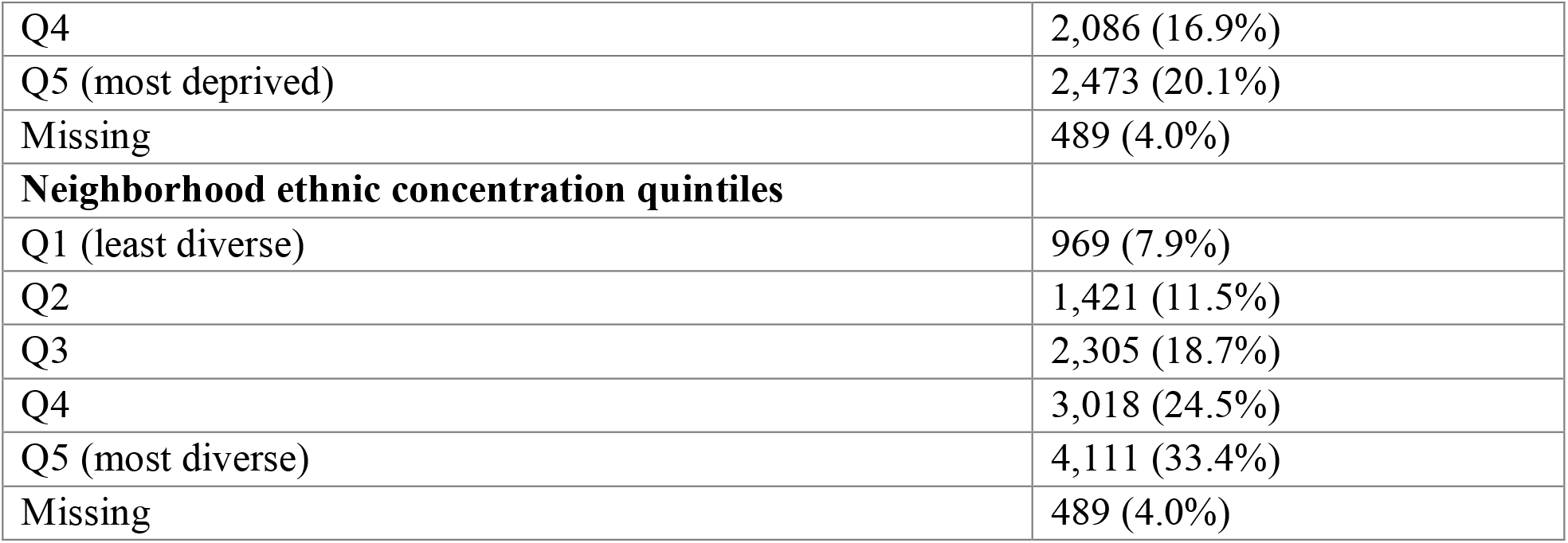
Characteristics of the study population.

| **All included children N = 12,313** | **N (%)** |
| --- | --- |
| **Birth year** |  |
| 2017 | 3,400 (27.6%) |
| 2018 | 3,412 (27.7%) |
| 2019 | 3,178 (25.8%) |
| 2020 | 2,323 (18.9%) |
| **Sex** |  |
| Female | 5,972 (48.5%) |
| Male | 6,341 (51.5%) |
| **Location** |  |
| Urban | 10,740 (87.2%) |
| Rural | 1,092 (8.9%) |
| Missing | 481 (3.9%) |
| **Neighborhood income quintiles** |  |
| Q1 (lowest income) | 2,757 (22.4%) |
| Q2 | 2,246 (18.2%) |
| Q3 | 2,224 (18.1%) |
| Q4 | 2,132 (17.3%) |
| Q5 (highest income) | 2,473 (20.1%) |
| Missing | 481 (3.9%) |
| **Neighborhood material deprivation quintiles** |  |
| Q1 (least deprived) | 2,559 (20.8%) |
| Q2 | 2,402 (19.5%) |
| Q3 | 2,304 (18.7%) |
| Q4 | 2,086 (16.9%) |
| Q5 (most deprived) | 2,473 (20.1%) |
| Missing | 489 (4.0%) |
| **Neighborhood ethnic concentration quintiles** |  |
| Q1 (least diverse) | 969 (7.9%) |
| Q2 | 1,421 (11.5%) |
| Q3 | 2,305 (18.7%) |
| Q4 | 3,018 (24.5%) |
| Q5 (most diverse) | 4,111 (33.4%) |
| Missing | 489 (4.0%) |

### UTD immunization coverage

At baseline (T1), the average overall UTD immunization coverage rate for all children under 2 years old was 71.0% (95% confidence interval (CI): 70.7, 71.3) (Figure 1). The UTD coverage rate decreased by 5.7% (95% CI: -6.2, -5.1) in the early pandemic period (T2). Later in the pandemic (T3), overall UTD coverage improved slightly but remained 4.1% (95% CI: -4.6, -3.6) below pre-pandemic levels.

**Figure 1.**
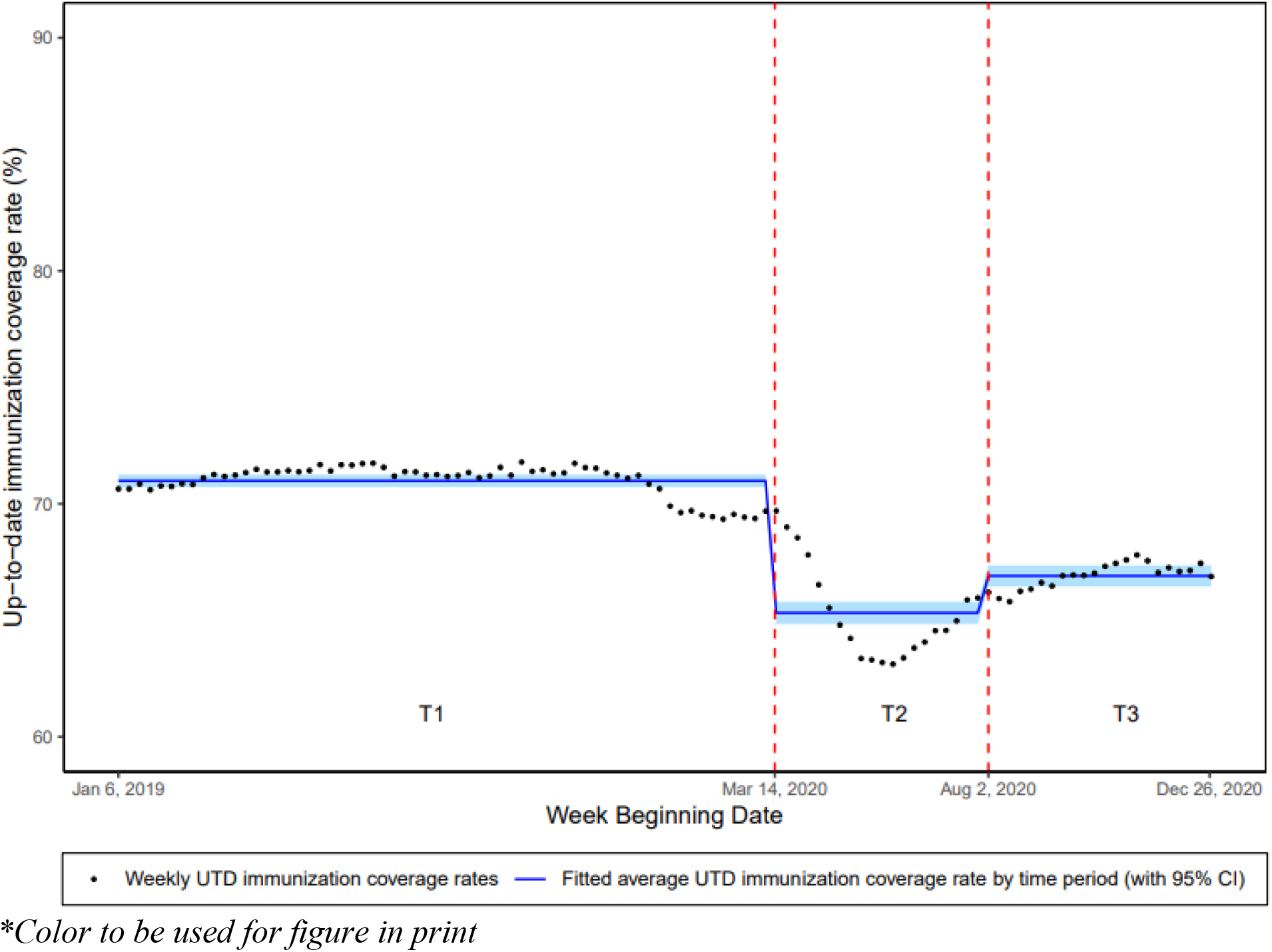
Overall up-to-date immunization coverage rate of children under 2 years old, from January 2019 to December 2020; weekly coverage rates and fitted average coverage rates by time period

When looking at UTD coverage for each specific vaccine, the average coverage rate in T1 was the lowest for varicella vaccine at 72.9% (95% CI: 72.5, 73.4) and was approximately 85% for the other vaccines (Figures 2a-b). The UTD coverage rates decreased for all vaccines in T2 when compared to T1, ranging from a 0.8% decrease (95% CI: -1.0, -0.6) for Pneu-C-13 to a 6.3% decrease (95% CI: -7.2, -5.4) for Var, and remained at similar reduced rates during T3. MMR vaccine coverage rate decreased by 1.7% (95% CI: -2.3, -1.2) in T2 and by 1.9% (95% CI: -2.4, - 1.4) in T3 when compared to the pre-pandemic period.

**Figure 2a.**
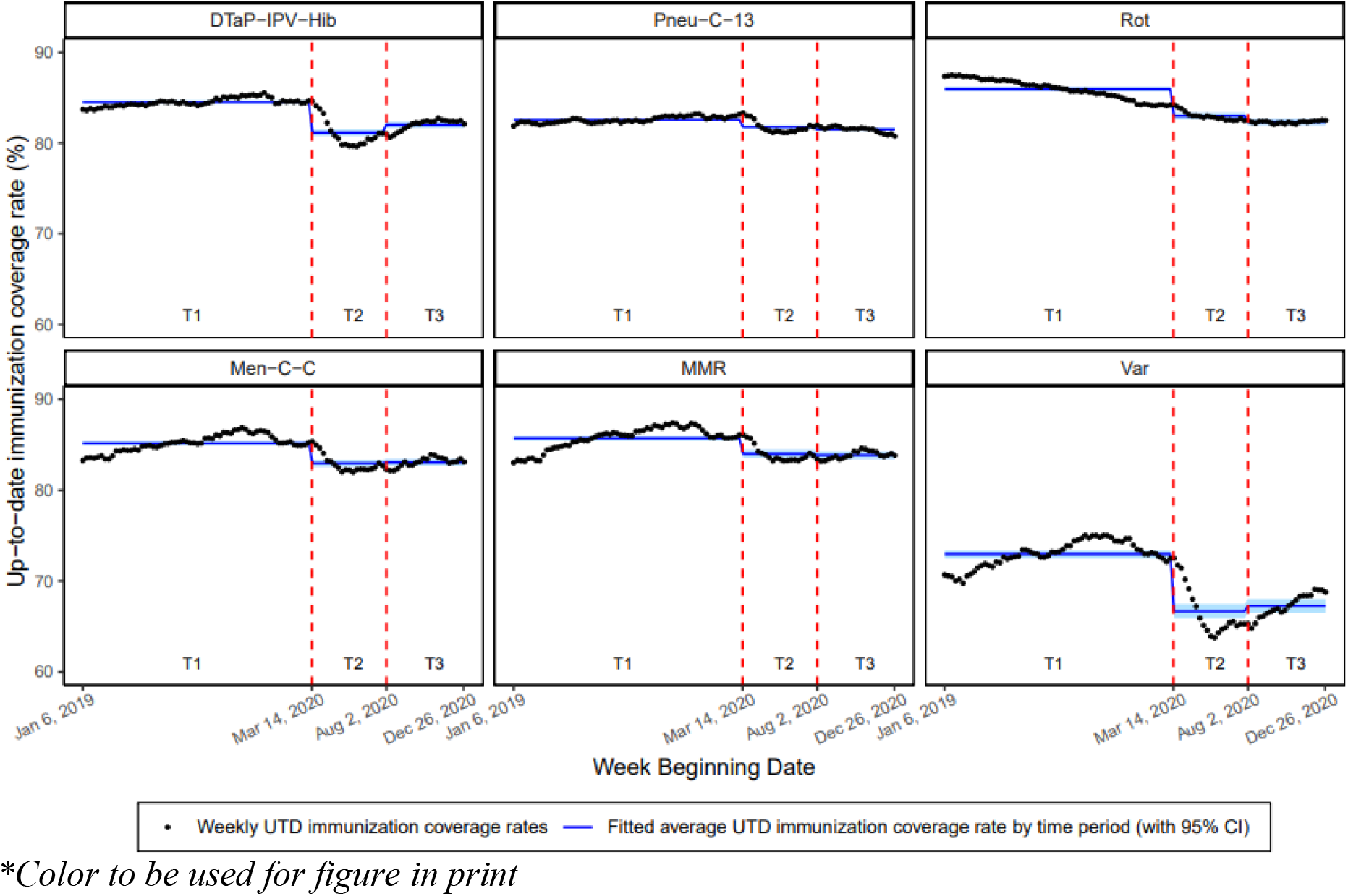
UTD immunization coverage rates for children under 2 years old by type of vaccine, from January 2019 to December 2020

**Figure 2b.**
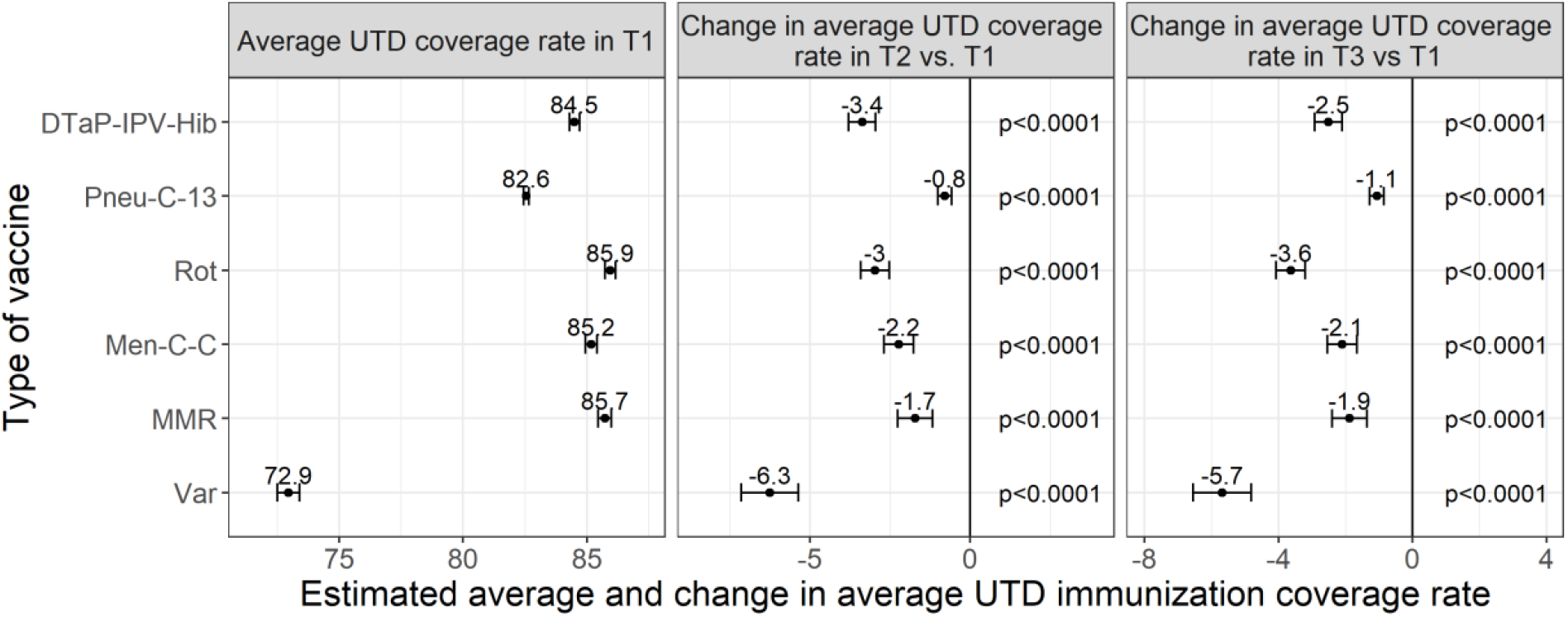
Average UTD immunization coverage rates by types of vacine in the pre-pandemic period (T1), and changes in the early pandemic period (T2) and in the later pandemic period (T3) compared to T1

### On-time immunization coverage

The baseline T1 average on-time immunization coverage rates were the highest for younger children and decreased progressively as children aged, from 88.3% (95% CI: 87.0, 89.5) for 2- month-old children down to 51.6% (95% CI: 49.8, 53.4) for 18-month-old children (Figures 3a-b). The largest impact from the pandemic was seen in older children, with drops in on-time coverage rates of 14.7% (95% CI: -18.7, -10.6) and 16.4% (95% CI: -20.0, -12.8) for 15- and 18- month-old children respectively during T2 compared to pre-pandemic period. These coverage rates improved in T3 but were still significantly lower than T1 rates. The 6- and 12-month-old children experienced decreases in on-time coverage rates during T2 of approximately 6.0%, but coverage rates recovered in T3 to pre-pandemic levels. On-time coverage rates for 2- and 4-month-old children were not significantly impacted by the pandemic throughout the study observation period.

**Figure 3a.**
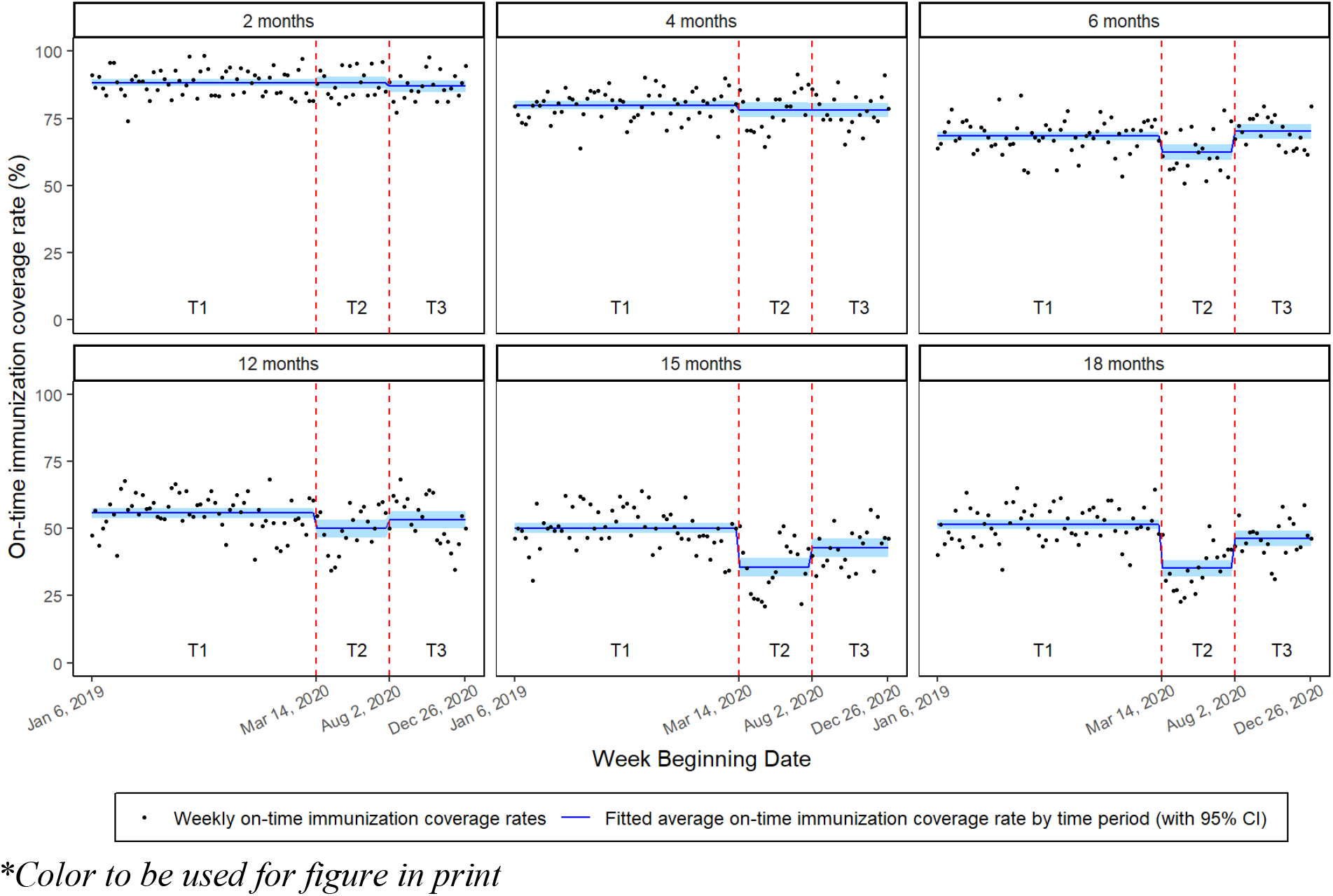
On-time immunization coverage rates by age milestone, from January 2019 to December 2020

**Figure 3b.**
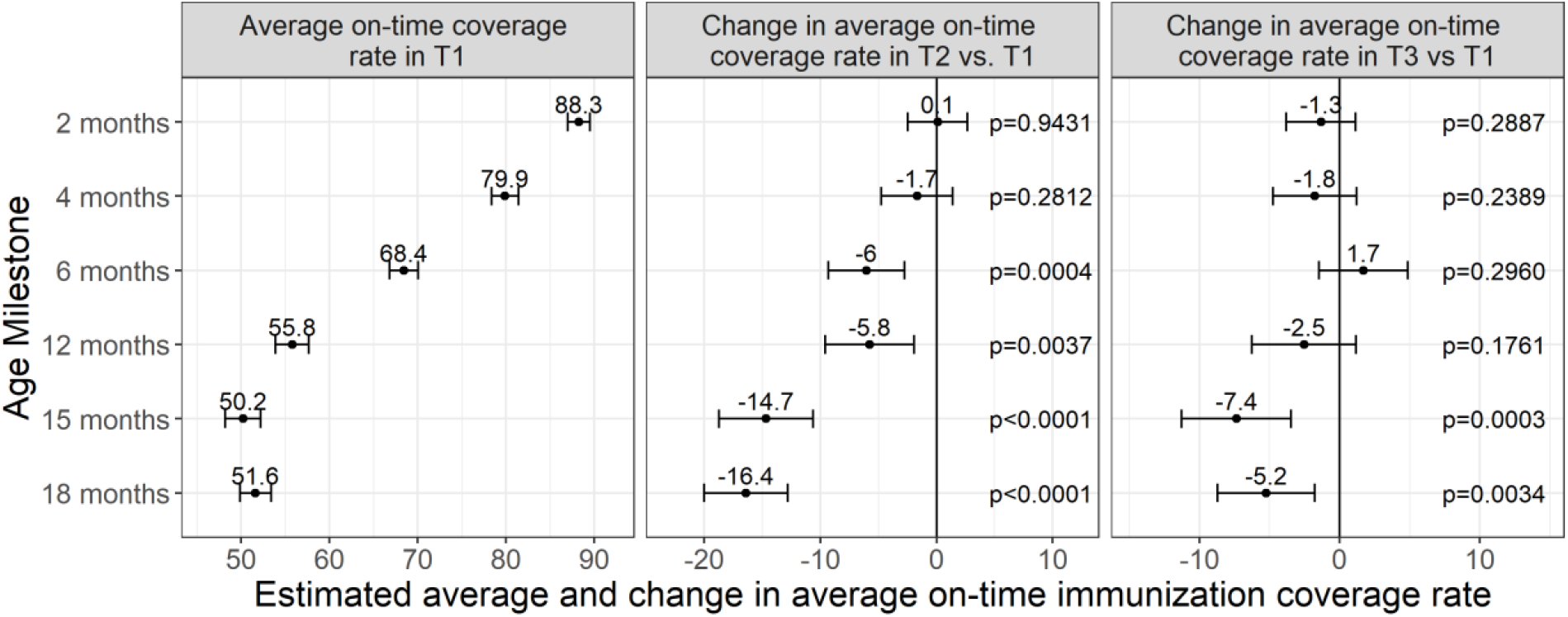
Average on-time immunization coverage rates by age milestone in the pre- pandemic period (T1), and change in the early pandemic period (T2) and in the later pandemic period (T3) compared to T1

When stratified by sociodemographic characteristics, baseline on-time coverage rates in T1 were not found to be statistically different by sex or by location across all age milestones, except in 4- month-old children living in urban areas who had a lower coverage rate than those living in rural areas (Figure 4). Neighborhood socioeconomic status measures, however, impacted baseline on-time coverage rates, with children living in neighborhoods in the lowest income, the most materially deprived and the most ethnically diverse quintiles showing significantly lower coverage rates than children from quintiles associated with higher SES, across all age milestones. When adjusted for these baseline differences, the variations in coverage rates over time (T2 vs T1 and T3 vs T1) were similar across all subgroups, including children of different sexes or from different locations and neighborhood SES, and across all age milestones. No specific subgroup was found to be differentially impacted by the pandemic by experiencing a significantly larger decrease in on-time immunization coverage.

**Figure 4.**
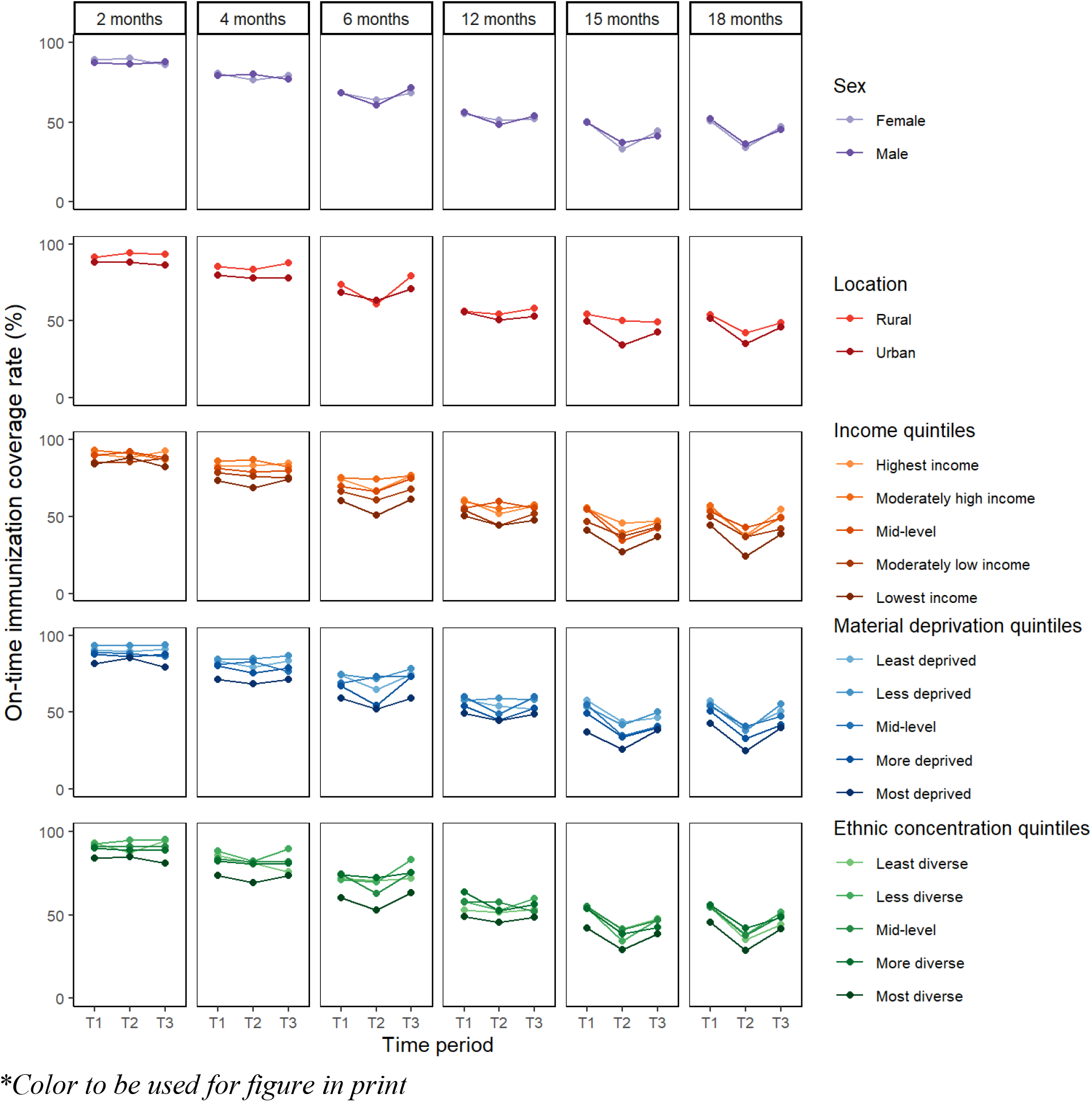
Average on-time immunization coverage rates by age milestone and sociodemographic characteristics, over the three time periods

## Discussion

In this retrospective study using EMR data from a network of family medicine clinics in the GTA, Ontario, we found that routine childhood immunization coverage rates in children under 2 years old decreased significantly in the early months of the COVID-19 pandemic, especially in children aged 15 and 18-months old. The coverage rates recovered in the second half of 2020 but remained lower overall compared to pre-pandemic rates. Our study provides more insight on which groups of children should be prioritized for immunization catch-up strategies and for targeted interventions to prevent immunization delays in future pandemic waves or other events that could be disruptive to the primary immunization schedule.

Similar to reports from other countries, we found that the impact of the pandemic and stay-at-home orders on immunization coverage rates was the most significant in the early period of the pandemic and only partially recovered subsequently [9-11, 38]. Some of this recovery may have been prompted by public messaging from official organizations that routine childhood immunization is an essential health service [9, 10]. In May 2020, Canada’s National Advisory Committee on Immunization recommended prioritizing primary immunization series up to 18-months old during the pandemic [18], which may have encouraged the increase in coverage rates seen in our results starting in June 2020. However, given lower rates persisted until the end of our observation period in December 2020, more education and support should be put forward for healthcare providers and parents [19].

Our finding that older (15- and 18-month-old) children have experienced a more significant drop in immunization coverage than younger children in the early pandemic period has been noted in other parts of the world [9-11]. Our study also found that the baseline on-time immunization coverage rate fell from around 88% for 2-month-old children to 51% at 18-months old. A similar trend was reported in the United States where the baseline vaccination coverage rate decreased from around 90% for 1-month-old children to 70% at 16-months old [11]. As children age, it is possible that there are more delays in attending routine visits as parents go back to working full-time [39]. Additionally, there may be less perceived infectious risks for older children, especially if staying home during lockdown, which may explain why we found that the varicella vaccine coverage was the lowest at baseline and decreased the most after the pandemic onset as it is given as a single vaccine at 15-months old. The suboptimal varicella vaccine coverage and the observed coverage gap where up to 50% of older infants are delayed in their immunizations leave many children vulnerable to VPDs, especially if attending daycare.

The MMR vaccine coverage is an often-studied outcome as measles is a highly transmissible VPD. Substantial drops in MMR vaccine coverage and administration in children have been observed in the early pandemic period in several countries [10, 11, 40]. In our study, we found that MMR vaccine UTD coverage for children under two dropped only by 1.7% in the early pandemic period and remained at the same reduced level during the rest of 2020. However, even small (2–5%) reductions in measles vaccination coverage have been projected to result in exponential increase in measles outbreak size [41]. Close monitoring of new cases of measles and other VPDs will therefore be important as lockdown measures and travel restrictions are lifted due to the potential risk of imported cases. In the United States, vaccine administrations plummeted across all racial and ethnic groups during the initial lockdown period, but recovery was lower in non-Hispanic Black children than in other groups during the reopening period [11]. In our study, we did not have patient-level ethnicity information, but we stratified our analyses by neighborhood ethnic concentration quintiles, in addition to income and material deprivation quintiles. Although the decrease in on-time coverage rates during the pandemic was found to be similar across all socioeconomic subgroups in our study, there was a significant difference in the baseline on-time coverage rates, with children from the lowest income, the most materially deprived and the most ethnically diverse quintiles having the lowest immunization coverage rates. These neighborhood quintiles have been associated with higher levels of marginalization and to health inequities in other studies [23]. Public health interventions and catch-up immunization programs should be preferentially implemented in these low income and ethnically diverse neighborhoods to provide equitable access to immunization services and improve coverage gaps [42, 43].

As COVID-19 number of cases and deaths continued to surge in subsequent waves and as vaccines against COVID-19 have been rolled out in Ontario in 2021, further studies are needed to trend the impact on childhood immunization coverage rates during 2021. Additional studies are also needed to better understand immunization coverage discrepancies between certain neighborhoods and potential association with other individual-, family- and health system-level factors, and to quantify the impact of the pandemic on other immunizations, such as school-based immunization programs.

## Limitations

The UTOPIAN data is limited to patients living mostly in the GTA, where there was a high number of COVID-19 cases and deaths, so our results may not be generalizable to other settings with lower COVID-19 incidence. Additionally, the patients in our study had access to family physicians working in clinical practices affiliated with the University of Toronto that likely had access to personal protective equipment and remained open to in-person appointments during the pandemic. Our results may therefore not be generalizable to children with limited access to primary care/family physicians, as these children may have experienced additional barriers to immunization services during the pandemic. The use of EMR data may lead to children being misclassified as not up-to-date or on-time in their immunizations if children received vaccines outside of their primary care practices or if their records are not adequately updated, which may have limited our analyses and led to an under-estimation of immunization coverage. However, primary care EMR data has been evaluated and shown to be a reliable source for childhood immunization coverage analyses in Ontario [31, 32, 36] as immunization services are mainly given in primary care clinics for infants and pre-school children. We still aimed to mitigate the possible impact of missing immunization records by only including patients with at least two visits with UTOPIAN family physicians to ensure continuity of care and exclude children who only saw the UTOPIAN-contributing family physician once (eg. walk-in visit, newborn visit, etc.) and completed their well-baby visits elsewhere.

## Conclusion

Our study found that childhood immunization coverage rates for children under 2 in Ontario, Canada were significantly decreased during the early period of the COVID-19 pandemic and only partially recovered during the rest of 2020. Implementation of public health and primary care interventions for providers and parents are needed to ensure adequate catch-up of delayed/missed immunizations to prevent potential outbreaks of vaccine-preventable diseases as these children enter daycare/kindergarten, especially in certain age and socioeconomic groups where baseline immunization coverage gaps have been identified.

## Data Availability

All data produced in the present study are available upon reasonable request to the authors

## Conflict of interest statement

The authors declare the following financial interests/personal relationships which may be considered as potential competing interests:

CJ, PPPR, JNF, MS, SZ and SKM are investigators on the operational research grant received from Pfizer Canada to conduct this investigator-led study. SKM has received honouraria for lectures from GlaxoSmithKline and was a member of an ad hoc advisory board for Pfizer Canada, both of which are unrelated to this study.

## Acknowledgement

We would like to acknowledge and thank Dr Sharon Burey for her support in the funding application of this study and Dr Babak Aliarzadeh for extracting the data from the UTOPIAN database and helping with the data cleaning process. Drs Catherine Ji and Karen Tu received Research Scholar Awards from the Department of Family and Community Medicine and the Rathlyn Foundation Primary Care EMR Research and Discovery Fund at the University of Toronto. Dr Ellen Stephenson was supported by a Fellowship award from the Canadian Institutes of Health Research.

## Funding statement

This work was supported by Pfizer Canada Competitive Grant Program [grant number 63478895]. The funder of the study had no role in study design, data collection, data analysis, data interpretation, writing of the report or decision to submit the article for publication.

## Contribution Statement

**Catherine Ji:** Conceptualization, Methodology, Data curation, Funding acquisition, Writing - original draft. **Pierre-Philippe Piché-Renaud:** Conceptualization, Methodology, Data curation, Funding acquisition, Writing - original draft. **Jemisha Apajee:** Methodology, Data curation, Formal analysis, Software, Writing - original draft. **Ellen Stephenson:** Methodology, Data curation, Formal analysis, Writing - review & editing. **Milena Forte**: Methodology, Writing - review & editing. **Jeremy N. Friedman:** Conceptualization, Funding acquisition, Writing - review & editing. **Michelle Science**: Conceptualization, Funding acquisition, Writing - review & editing. **Stanley Zlotkin**: Conceptualization, Funding acquisition, Writing - review & editing. **Shaun K. Morris:** Conceptualization, Methodology, Funding acquisition, Writing - review & editing. **Karen Tu**: Conceptualization, Methodology, Data curation, Writing - review & editing. All authors have approved the final version of this manuscript.

## Notes

### Author Declarations

REB of University Health Network gave ethical approval for this work. REB of University of Toronto Health Sciences gave ethical approval for this work.

## References

[1] World Health Organization. WHO and UNICEF warn of a decline in vaccinations during COVID-19. 2020.

[2] Chanchlani N, Buchanan F, Gill PJ. Addressing the indirect effects of COVID-19 on the health of children and young people. CMAJ. 2020;192:E921–E7.

[3] Stephenson E, Butt DA, Gronsbell J, Ji C, O’Neill B, Crampton N, et al. Changes in the top 25 reasons for primary care visits during the COVID-19 pandemic in a high-COVID region of Canada. PloS one. 2021;16:e0255992.

[4] Santoli JM LM, DeSilva MB, et al. Effects of the COVID-19 pandemic on routine pediatric vaccine ordering and administration—United States, 2020. MMWR Morbidity and mortality weekly report. 2020;69:591–3.

[5] Harris RC, Chen Y, Côte P, Ardillon A, Nievera MC, Ong-Lim A, et al. Impact of COVID-19 on routine immunisation in South-East Asia and Western Pacific: Disruptions and solutions. The Lancet Regional Health-Western Pacific. 2021:100140.

[6] Bramer CA KL, Swanson R, et al. Decline in Child Vaccination Coverage During the COVID-19 Pandemic — Michigan Care Improvement Registry, May 2016–May 2020. MMWR Morb Mortal Wkly Rep. 2020;69:630–1.

[7] World Health Organization. Special feature: immunization and COVID-19. 2020.

[8] Lassi ZS, Naseem R, Salam RA, Siddiqui F, Das JK. The impact of the COVID-19 pandemic on immunization campaigns and programs: a systematic review. International journal of environmental research and public health. 2021;18:988.

[9] Aizawa Y, Katsuta T, Sakiyama H, Tanaka-Taya K, Moriuchi H, Saitoh A. Changes in childhood vaccination during the coronavirus disease 2019 pandemic in Japan. Vaccine. 2021;39:4006–12.

[10] McDonald HI, Tessier E, White JM, Woodruff M, Knowles C, Bates C, et al. Early impact of the coronavirus disease (COVID-19) pandemic and physical distancing measures on routine childhood vaccinations in England, January to April 2020. Eurosurveillance. 2020;25:2000848.

[11] Ackerson BK, Sy LS, Glenn SC, Qian L, Park CH, Riewerts RJ, et al. Pediatric Vaccination During the COVID-19 Pandemic. Pediatrics. 2021.

[12] Causey K, Fullman N, Sorensen RJ, Galles NC, Zheng P, Aravkin A, et al. Estimating global and regional disruptions to routine childhood vaccine coverage during the COVID-19 pandemic in 2020: a modelling study. The Lancet. 2021.

[13] Mulholland K, Kretsinger K, Wondwossen L, Crowcroft N. Action needed now to prevent further increases in measles and measles deaths in the coming years. The Lancet. 2020;396:1782–4.

[14] Durrheim DN, Andrus JK, Tabassum S, Bashour H, Githanga D, Pfaff G. A dangerous measles future looms beyond the COVID-19 pandemic. Nature Medicine. 2021;27:360–1.

[15] Rana MS, Alam MM, Ikram A, Salman M, Mere MO, Usman M, et al. Emergence of measles during the COVID-19 pandemic threatens Pakistan’s children and the wider region. Nature Medicine. 2021:1–2.

[16] World Health Organization. Immunization as an essential health service: guiding principles for immunization activities during the COVID-19 pandemic and other times of severe disruption, 1 November 2020. 2020.

[17] Centers for Disease Control and Prevention. Catch Up on Well-Child Visits and Recommended Vaccinations. 2021.

[18] Public Health Agency of Canada - National Advisory Committee on Immunization. Interim guidance on continuity of immunization programs during the COVID-19 pandemic. 2020.

[19] Piché-Renaud P-P, Ji C, Farrar DS, Friedman JN, Science M, Kitai I, et al. Impact of the COVID-19 pandemic on the provision of routine childhood immunizations in Ontario, Canada. Vaccine. 2021;39:4373–82.

[20] Sell H, Assi A, Driedger M, Dubé E, Gagneur A, Meyer SB, et al. Continuity of routine immunization programs in Canada during the COVID-19 pandemic. medRxiv. 2021.

[21] University of Toronto. UTOPIAN Data Safe Haven. 2021.

[22] Statistics Canada. Postal Code OM Conversion File Plus (PCCF+). 2021.

[23] Matheson F, Van Ingen T. Ontario marginalization index user guide. Ontario Agency for Health Protection and Promotion (Public Health Ontario). 2016;2018:23.

[24] Matheson FI, Dunn JR, Smith KL, Moineddin R, Glazier RH. Development of the Canadian Marginalization Index: a new tool for the study of inequality. Canadian Journal of Public Health/Revue Canadienne de Sante’e Publique. 2012:S12–S6.

[25] Statistics Canada. Population estimates, quarterly. 2021.

[26] Government of Ontario. Ontario Enacts Declaration of Emergency to Protect the Public. 2020.

[27] The Canadian Press. Toronto, Peel Region officially enter Stage 3 of Ontario’s reopening plan. 2020.

[28] Government of Ontario. All Ontario: Case numbers and spread. 2021.

[29] Government of Canada. Coronavirus disease (COVID-19): Outbreak update. [Cited 2020 August 18]; Retrieved from: https://www.canada.ca/en/public-health/services/diseases/2019-novel-coronavirus-infection.html..

[30] Wilson SE, Wilton AS, Young J, Candido E, Bunko A, Buchan SA, et al. Assessing the completeness of infant and childhood immunizations within a provincial registry populated by parental reporting: A study using linked databases in Ontario, Canada. Vaccine. 2020;38:5223–30.

[31] Wilson SE, Chung H, Schwartz KL, Guttmann A, Deeks SL, Kwong JC, et al. Rotavirus vaccine coverage and factors associated with uptake using linked data: Ontario, Canada. PloS one. 2018;13:e0192809.

[32] Schwartz KL, Tu K, Wing L, Campitelli MA, Crowcroft NS, Deeks SL, et al. Validation of infant immunization billing codes in administrative data. Human vaccines & immunotherapeutics. 2015;11:1840–7.

[33] Government of Ontario. Ontario’s Routine Immunization Schedule

[34] Toronto Public Health. Change in Rotavirus Vaccine. 2018.

[35] MacDonald SE, Russell ML, Liu XC, Simmonds KA, Lorenzetti DL, Sharpe H, et al. Are we speaking the same language? An argument for the consistent use of terminology and definitions for childhood vaccination indicators. Human vaccines & immunotherapeutics. 2019;15:740–7.

[36] Wilson SE, Quach S, MacDonald SE, Naus M, Deeks SL, Crowcroft NS, et al. Methods used for immunization coverage assessment in Canada, a Canadian Immunization Research Network (CIRN) study. Human vaccines & immunotherapeutics. 2017;13:1928–36.

[37] Rourke L. Rourke Baby Record.

[38] Murthy BP, Zell E, Kirtland K, Jones-Jack N, Harris L, Sprague C, et al. Impact of the COVID-19 Pandemic on Administration of Selected Routine Childhood and Adolescent Vaccinations—10 US Jurisdictions, March–September 2020. Morbidity and Mortality Weekly Report. 2021;70:840.

[39] Berger LM, Hill J, Waldfogel J. Maternity leave, early maternal employment and child health and development in the US. The Economic Journal. 2005;115:F29–F47.

[40] Middeldorp M, van Lier A, van der Maas N, Veldhuijzen I, Freudenburg W, van Sorge NM, et al. Short term impact of the COVID-19 pandemic on incidence of vaccine preventable diseases and participation in routine infant vaccinations in the Netherlands in the period March-September 2020. Vaccine. 2021;39:1039–43.

[41] Phadke VK, Bednarczyk RA, Omer SB. Vaccine refusal and measles outbreaks in the US. Jama. 2020;324:1344–5.

[42] Vann JCJ, Jacobson RM, Coyne-Beasley T, Asafu-Adjei JK, Szilagyi PG. Patient reminder and recall interventions to improve immunization rates. Cochrane Database of Systematic Reviews. 2018.

[43] Machado AA, Edwards SA, Mueller M, Saini V. Effective interventions to increase routine childhood immunization coverage in low socioeconomic status communities in developed countries: A systematic review and critical appraisal of peer-reviewed literature. Vaccine. 2021.

